# Risks Associated with Animal-Assisted Intervention Programs: A Literature Review

**DOI:** 10.1101/2020.02.19.20025130

**Authors:** Kathryn R. Dalton, Kaitlin B. Waite, Kathy Ruble, Karen C. Carroll, Alexandra DeLone, Pam Frankenfield, James A. Serpell, Roland J. Thorpe, Daniel O. Morris, Jacqueline Agnew, Ronald C. Rubenstein, Meghan F. Davis

## Abstract

The benefits of animal-assisted interventions (AAI), to utilize companion animals as an adjunctive treatment modality, is well-established and a burgeoning research field. However, few studies have evaluated the potential hazards of these programs, such as the potential for therapy animals to transfer hospital-associated pathogens between individuals and the hospital environment. Here we review the current literature on the possible risks of hospital-based AAI programs, including zoonotic pathogen transmission. We identified twenty-nine articles encompassing reviews of infection control guidelines and epidemiological studies on zoonotic pathogen prevalence in AAI. We observed substantial heterogeneity in infection control practices among hospital AAI programs. Few data confirmed pathogen transmission between therapy animals and patients. Given AAI’s known benefits, we recommend that future research utilize a One Health framework to evaluate microbial dynamics among therapy animals, patients, and hospital environments. This framework may best promote safe practices to ensure the sustainability of these valuable AAI programs.

**Highlights:**

- Despite the many benefits of animal-assisted interventions (AAI) for patients, there is a risk of therapy animals becoming vectors of hospital pathogens.
- There is an absence of literature on transmission of hospital pathogens between patients and therapy animals during an AAI session.
- More research is needed to improve the safety and utilization of this important adjunctive therapy.

## 1. Introduction

The emotional benefits of human-companion animal relationships are well established in the scientific literature (Serpell, 1996). This concept has extended into the development of animal-assisted interventions (AAI), in which visiting animals participate as an adjunctive treatment in holistic patient care. AAI programs are increasingly popular in various healthcare settings and utilized for patients with widely diverse conditions, including mental health disorders and cancer. Research into the benefits of AAI continues to expand, with the many advantages of these programs supported by numerous epidemiological studies and meta-analyses that standardize and integrate these findings. These data support the hypothesis that AAI programs reduce patient stress, pain, and anxiety levels when incorporated into patients’ treatment plans (Bert et al., 2016; Kamioka et al., 2014; Lundqvist et al., 2017; Maujean et al., 2015; Serpell et al., 2017).

However, infection control is a persistent problem in healthcare settings, both in routine care and in the use of complementary therapies. Similar to known fomites in hospitals, such as door handles and clinicians’ stethoscopes (Haun et al., 2016), therapy animals may unwittingly serve as mechanical vectors of hospital-associated pathogens, and contribute to the transmission of these pathogens between patients, or otherwise within the hospital environment. Patients can experience different levels of animal exposure from petting and licking, which can result in contamination of both the patient and the animal, thereby providing the opportunity for the spread of microorganisms (Lefebvre & Weese, 2009). Therapy animals also have the potential to introduce zoonotic pathogens directly into the hospital environment, for example, via the consumption of contaminated foods (Lefebvre et al., 2008b). Contamination by a pathogen could potentially lead to pathogen replication and stable colonization; this is concerning not only for the possible risk of progression to infection, but also for the risk that the therapy animal may serve as a reservoir and spread these pathogens to the home and larger community (Enoch et al., 2005). Such perceptions of potential infection control challenges and resulting harm could limit the use of AAI programs and detract from their employment as a valid and valuable adjunctive therapy for patients.

This review examines the current literature that focuses on potential hazards associated with hospital-based AAI therapy programs. We assessed both the breadth and quality of existing literature regarding infection control in AAI programs; these are discussed in the context of known and hypothetical pathways of microbial transmission. By identifying knowledge gaps, we provide focus for future research efforts and intervention strategies that will ultimately promote the sustainability of these AAI programs.

## 2. Methods

### 2.1 Search Strategy

This review utilized a more flexible search strategy in order to optimize capture of the peer-reviewed literature related to the risk of animal-assisted therapy. Multiple search approaches and terminology were employed to capture existing evidence relating to animal-assisted interventions for patients as a whole. Several unique terms can apply to AAI, such as animal-assisted therapy, animal-assisted activities, or pet therapy, therefore the search strategy was intentionally broad.

The literature search on risks of animal use in hospitals was carried out using the following databases: PubMed, Scopus, Embase, Web of Science, CINAHL, and Cochrane Trials. The search was completed concurrently and independently by two of the authors (KRD, KBW), and the search strategy was framed using PICO (Population, Intervention/Exposure, Comparators, Outcomes) terms (Miller & Forrest, 2001). The Population was identified as healthcare-based AAI programs using any therapy animals, not just canines. The Intervention/Exposure and Comparators were kept flexible and were dependent on study design. The Outcomes were any potential hazards associated with AAI, particularly infectious disease, microbial, or biological risks. Study designs accepted for review remained flexible and included original epidemiological research, literature reviews, commentaries, and case-reports.

### 2.2 Search Terms

In collaboration with a librarian, we performed a systematic search using the terms listed below on the respective databases; search terms were adjusted according to individual database terminologies, and searches were restricted to title/abstract. We used the following search strategy for the PubMed database: animal assist* OR pet assist* OR dog assist* OR pet therap* OR dog therap* OR animal therap* OR “animal facilitated” OR “pet facilitated” OR “therapeutic animal” OR “therapeutic animals” OR “therapeutic canine” OR “therapeutic canines” OR “therapeutic dog” OR “therapeutic dogs” OR [Animal Assisted Intervention MeSH Term]. Similar keywords were used to conduct searches within the other selected databases.

### 2.3 Inclusion and Exclusion Criteria

The articles identified from this broad search were then individually and independently screened by two of the authors (KRD, KBW), based on the title and abstract, for inclusion based on the following criteria:

▪ Did the article explain possible complications or hazards to either therapy animals or patients that can occur during a hospital AAI therapy session?
▪ Did the article describe an epidemiological study demonstrating the risk of animals within health care environments?
▪ Did the article provide novel commentary on current guidelines, or recommend new guidelines, for reducing associated risks of animals within healthcare environments?

Articles that did not address any of the above criteria, or written in a language other than English, were excluded. Eligible studies underwent full-text review to further confirm eligibility (by KRD & KBW, arbiter MFD). After full-text review, references were examined to look for additional relevant articles that fit the inclusion criteria. We then extracted data from the selected studies on the research aims, study design, study population, exposure characteristics, type of intervention (if any), reported outcomes, and results. These data were then synthesized by study goals and outcomes.

## 3. Results

### 3.1 Search Outcomes

The initial database search returned a total of 5480 unique results (maximum number of returned articles from Embase), as shown in the flow diagram in **Figure 1**. After title and abstract screening of these articles, 110 were deemed potentially relevant based on the inclusion criteria. The remaining 5370 articles did not meet our prespecified criteria for inclusion, most commonly because the excluded articles evaluated the benefits of AAI programs on patient care. Upon full-text review of the 110 potentially relevant articles, 86 articles were removed because they did not satisfy the inclusion criteria. An additional five articles were added after reviewing the reference lists of the remaining included papers. These five articles were not found in the initial database search because they were either 1) not located in the selected databases or 2) had improperly labeled keywords. A summary of the final 29 total articles reviewed can be found in **Tables 1 and 2**. Thirteen articles were reviews or commentaries of current AAI guidelines that refer to therapy animals in healthcare settings, and sixteen articles were data-acquiring or epidemiological studies (6 cohort studies, 5 cross-sectional studies, 4 case reports, and 1 ecological study). Most studies focused on therapy animals broadly or therapy dogs exclusively, but three studies included cats (Boyle et al., 2019; Coughlan et al., 2010; Sillery et al., 2004).

**Table 1.**
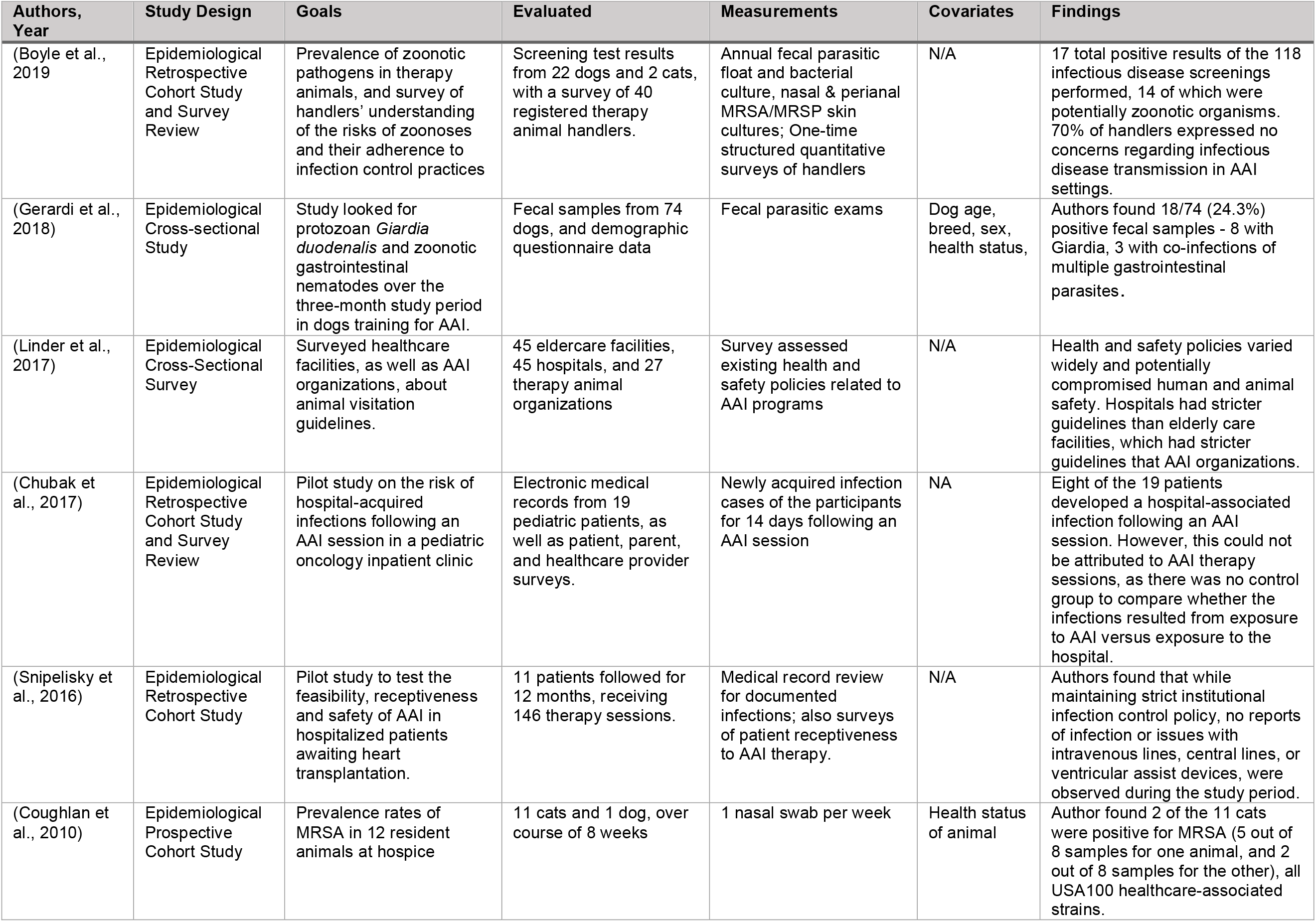
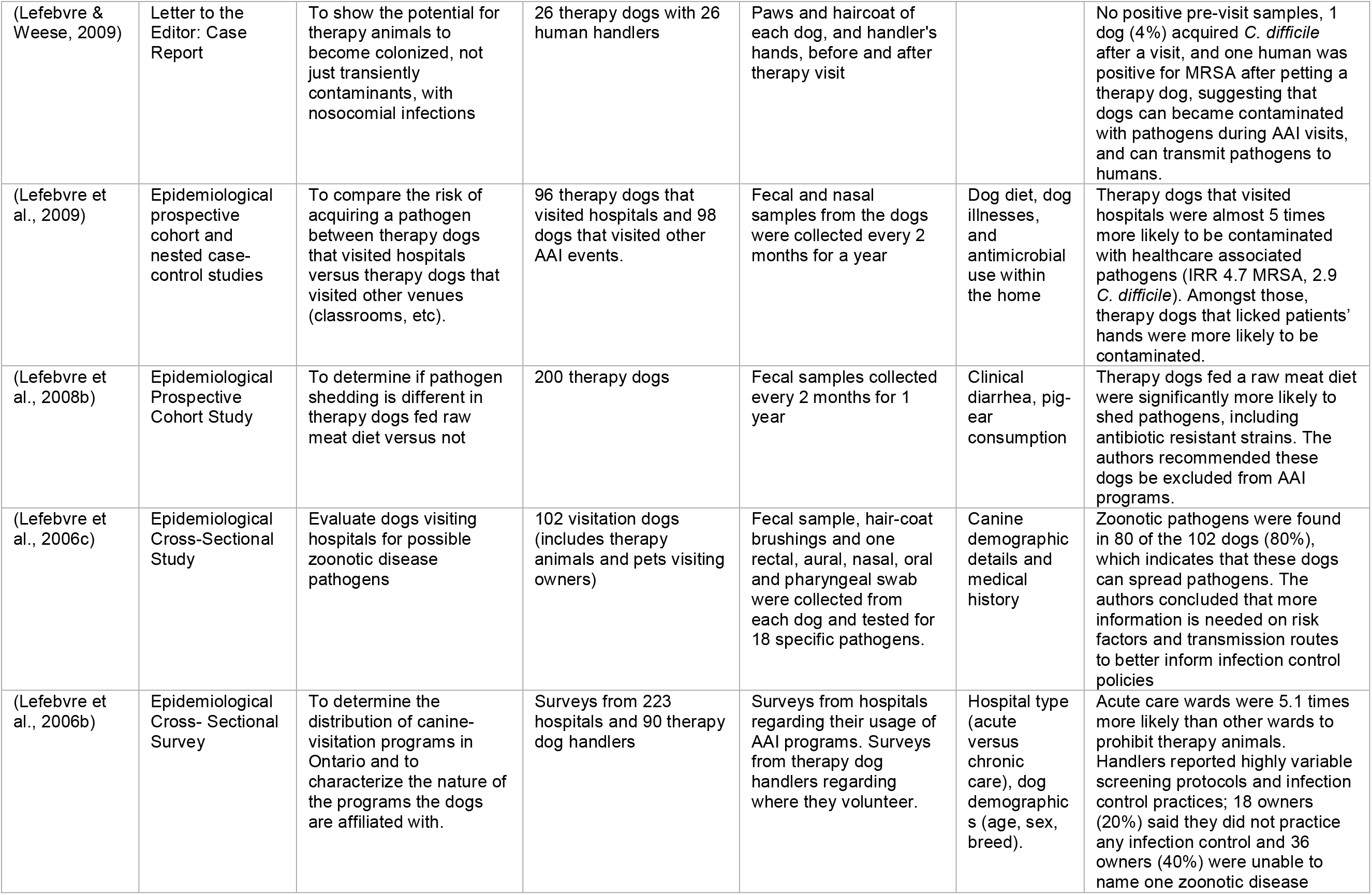
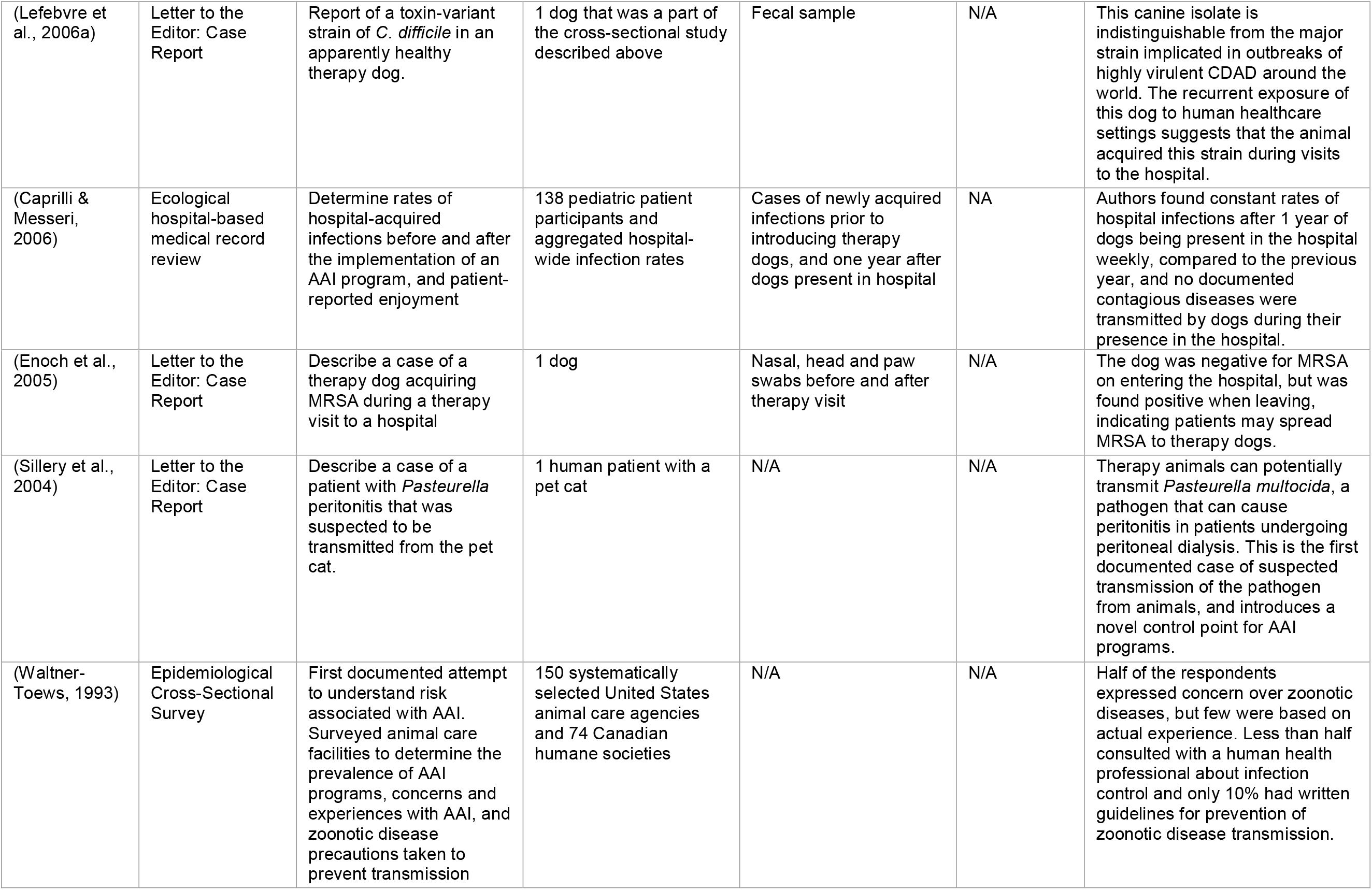
**Overview of the Articles Examined: Epidemiological Studies**, listed by type and chronologically

| Authors, Year | Study Design | Goals | Evaluated | Measurements | Covariates | Findings |
| --- | --- | --- | --- | --- | --- | --- |
| (Boyle et al., 2019) | Epidemiological Retrospective Cohort Study and Survey Review | Prevalence of zoonotic pathogens in therapy animals, and survey of handlers' understanding of the risks of zoonoses and their adherence to infection control practices | Screening test results from 22 dogs and 2 cats, with a survey of 40 registered therapy animal handlers. | Annual fecal parasitic float and bacterial culture, nasal & perianal MRSA/MRSP skin cultures; One-time structured quantitative surveys of handlers | N/A | 17 total positive results of the 118 infectious disease screenings performed, 14 of which were potentially zoonotic organisms. 70% of handlers expressed no concerns regarding infectious disease transmission in AAI settings. |
| (Gerardi et al., 2018) | Epidemiological Cross-sectional Study | Study looked for protozoan *Giardia duodenalis* and zoonotic gastrointestinal nematodes over the three-month study period in dogs training for AAI. | Fecal samples from 74 dogs, and demographic questionnaire data | Fecal parasitic exams | Dog age, breed, sex, health status, | Authors found 18/74 (24.3%) positive fecal samples - 8 with *Giardia*, 3 with co-infections of multiple gastrointestinal parasites. |
| (Linder et al., 2017) | Epidemiological Cross-Sectional Survey | Surveyed healthcare facilities, as well as AAI organizations, about animal visitation guidelines. | 45 eldercare facilities, 45 hospitals, and 27 therapy animal organizations | Survey assessed existing health and safety policies related to AAI programs | N/A | Health and safety policies varied widely and potentially compromised human and animal safety. Hospitals had stricter guidelines than elderly care facilities, which had stricter guidelines than AAI organizations. |
| (Chubak et al., 2017) | Epidemiological Retrospective Cohort Study and Survey Review | Pilot study on the risk of hospital-acquired infections following an AAI session in a pediatric oncology inpatient clinic | Electronic medical records from 19 pediatric patients, as well as patient, parent, and healthcare provider surveys. | Newly acquired infection cases of the participants for 14 days following an AAI session | NA | Eight of the 19 patients developed a hospital-associated infection following an AAI session. However, this could not be attributed to AAI therapy sessions, as there was no control group to compare whether the infections resulted from exposure to AAI versus exposure to the hospital. |
| (Snipelisky et al., 2016) | Epidemiological Retrospective Cohort Study | Pilot study to test the feasibility, receptiveness and safety of AAI in hospitalized patients awaiting heart transplantation. | 11 patients followed for 12 months, receiving 146 therapy sessions. | Medical record review for documented infections; also surveys of patient receptiveness to AAI therapy. | N/A | Authors found that while maintaining strict institutional infection control policy, no reports of infection or issues with intravenous lines, central lines, or ventricular assist devices, were observed during the study period. |
| (Coughlan et al., 2010) | Epidemiological Prospective Cohort Study | Prevalence rates of MRSA in 12 resident animals at hospice | 11 cats and 1 dog, over course of 8 weeks | 1 nasal swab per week | Health status of animal | Author found 2 of the 11 cats were positive for MRSA (5 out of 8 samples for one animal, and 2 out of 8 samples for the other), all USA100 healthcare-associated strains. |
| (Lefebvre & Weese, 2009) | Letter to the Editor: Case Report | To show the potential for therapy animals to become colonized, not just transiently contaminants, with nosocomial infections | 26 therapy dogs with 26 human handlers | Paws and haircoat of each dog, and handler's hands, before and after therapy visit |  | No positive pre-visit samples, 1 dog (4%) acquired *C. difficile* after a visit, and one human was positive for MRSA after petting a therapy dog, suggesting that dogs can become contaminated with pathogens during AAI visits, and can transmit pathogens to humans. |
| (Lefebvre et al., 2009) | Epidemiological prospective cohort and nested case-control studies | To compare the risk of acquiring a pathogen between therapy dogs that visited hospitals versus therapy dogs that visited other venues (classrooms, etc). | 96 therapy dogs that visited hospitals and 98 dogs that visited other AAI events. | Fecal and nasal samples from the dogs were collected every 2 months for a year | Dog diet, dog illnesses, and antimicrobial use within the home | Therapy dogs that visited hospitals were almost 5 times more likely to be contaminated with healthcare associated pathogens (IRR 4.7 MRSA, 2.9 *C. difficile*). Amongst those, therapy dogs that licked patients' hands were more likely to be contaminated. |
| (Lefebvre et al., 2008b) | Epidemiological Prospective Cohort Study | To determine if pathogen shedding is different in therapy dogs fed raw meat diet versus not | 200 therapy dogs | Fecal samples collected every 2 months for 1 year | Clinical diarrhea, pig-ear consumption | Therapy dogs fed a raw meat diet were significantly more likely to shed pathogens, including antibiotic resistant strains. The authors recommended these dogs be excluded from AAI programs. |
| (Lefebvre et al., 2006c) | Epidemiological Cross-Sectional Study | Evaluate dogs visiting hospitals for possible zoonotic disease pathogens | 102 visitation dogs (includes therapy animals and pets visiting owners) | Fecal sample, hair-coat brushings and one rectal, aural, nasal, oral and pharyngeal swab were collected from each dog and tested for 18 specific pathogens. | Canine demographic details and medical history | Zoonotic pathogens were found in 80 of the 102 dogs (80%), which indicates that these dogs can spread pathogens. The authors concluded that more information is needed on risk factors and transmission routes to better inform infection control policies |
| (Lefebvre et al., 2006b) | Epidemiological Cross- Sectional Survey | To determine the distribution of canine-visitation programs in Ontario and to characterize the nature of the programs the dogs are affiliated with. | Surveys from 223 hospitals and 90 therapy dog handlers | Surveys from hospitals regarding their usage of AAI programs. Surveys from therapy dog handlers regarding where they volunteer. | Hospital type (acute versus chronic care), dog demographics (age, sex, breed). | Acute care wards were 5.1 times more likely than other wards to prohibit therapy animals. Handlers reported highly variable screening protocols and infection control practices; 18 owners (20%) said they did not practice any infection control and 36 owners (40%) were unable to name one zoonotic disease |
| (Lefebvre et al., 2006a) | Letter to the Editor: Case Report | Report of a toxin-variant strain of *C. difficile* in an apparently healthy therapy dog. | 1 dog that was a part of the cross-sectional study described above | Fecal sample | N/A | This canine isolate is indistinguishable from the major strain implicated in outbreaks of highly virulent CDAD around the world. The recurrent exposure of this dog to human healthcare settings suggests that the animal acquired this strain during visits to the hospital. |
| (Caprilli & Messeri, 2006) | Ecological hospital-based medical record review | Determine rates of hospital-acquired infections before and after the implementation of an AAI program, and patient-reported enjoyment | 138 pediatric patient participants and aggregated hospital-wide infection rates | Cases of newly acquired infections prior to introducing therapy dogs, and one year after dogs present in hospital | NA | Authors found constant rates of hospital infections after 1 year of dogs being present in the hospital weekly, compared to the previous year, and no documented contagious diseases were transmitted by dogs during their presence in the hospital. |
| (Enoch et al., 2005) | Letter to the Editor: Case Report | Describe a case of a therapy dog acquiring MRSA during a therapy visit to a hospital | 1 dog | Nasal, head and paw swabs before and after therapy visit | N/A | The dog was negative for MRSA on entering the hospital, but was found positive when leaving, indicating patients may spread MRSA to therapy dogs. |
| (Sillery et al., 2004) | Letter to the Editor: Case Report | Describe a case of a patient with *Pasteurella* peritonitis that was suspected to be transmitted from the pet cat. | 1 human patient with a pet cat | N/A | N/A | Therapy animals can potentially transmit *Pasteurella multocida*, a pathogen that can cause peritonitis in patients undergoing peritoneal dialysis. This is the first documented case of suspected transmission of the pathogen from animals, and introduces a novel control point for AAI programs. |
| (Waltner-Toews, 1993) | Epidemiological Cross-Sectional Survey | First documented attempt to understand risk associated with AAI. Surveyed animal care facilities to determine the prevalence of AAI programs, concerns and experiences with AAI, and zoonotic disease precautions taken to prevent transmission | 150 systematically selected United States animal care agencies and 74 Canadian humane societies | N/A | N/A | Half of the respondents expressed concern over zoonotic diseases, but few were based on actual experience. Less than half consulted with a human health professional about infection control and only 10% had written guidelines for prevention of zoonotic disease transmission. |

**Table 2.** **Overview of the Articles Examined: Reviews, Guidelines, and Commentaries**, listed by type and chronologically

| First Author, Year | Study Design | Goals | Evaluated | Measurements | Covariates | Findings |
| --- | --- | --- | --- | --- | --- | --- |
| (Bert et al., 2016) | Systematic Review | Review current literature of positive clinical outcomes and negative risk to patients from therapy animals | 11 papers looking at the risk of therapy animals, which include both epidemiological studies and protocol guidelines. | N/A | N/A | Concluded AAI for hospitalized patients useful and safe for a wide range of diseases |
| (Chalmers & Dell, 2016) | Commentary | Applying One Health principles to decrease risk in therapy dog programs and further research | Did not include number of papers formally reviewed | N/A | N/A | Author gives a framework for studying therapy programs in the animal-human-environment interface. |
| (Hardin et al., 2016) | Commentary | Describe implementation of a pet therapy program that includes guidelines for the prevention of transmitted infections. | Did not include number of papers formally reviewed | N/A | N/A | Guidelines were in place in a hospital for sixteen years with no documented cases of disease transmission, supporting that a pet therapy program can be put into place safely with proper regulation |
| (Cimolai, 2015) | Letter to the Editor: Brief Review | Short review of current studies/case reports of zoonotic infections from pets | Did not include number of papers formally reviewed | N/A | N/A | Author concludes that therapy programs do provide opportunities for patients to become exposed to zoonotic infections and requires strict infection control policies, not a relaxation of guidelines. |
| (Murthy et al., 2015), Society of Healthcare Epidemiology of America (SHEA) Writing Group | Commentary | Provide general guidance to the medical community regarding management of animals in healthcare, specifically in terms of hazard reduction. | Did not include number of papers formally reviewed | N/A | N/A | Created guidelines for animal-assisted therapies, service animals, research animals, and personal pet visitation. Also recommends additional research be performed to better understand the risks and benefits of allowing animals in the healthcare setting for specific purposes |
| (Snipelisky & Burton, 2014) | Review | Review current published information regarding the efficacy of AAI in the inpatient population, and to review safety concerns associated with AAI. | Reviewed 44 articles (26 clinical studies, 15 review articles, 1 case report and 2 letters to the editor). Five studies addressed infection concerns. | N/A | N/A | The authors' review of the literature showed that, in the inpatient setting, AAI is an effective therapy among patients of all ages and with various medical problems and is safe, with no transmitted infections reported. Found only 5 studies that addressed infection concerns in the inpatient setting. |
| (Silveira et al., 2011) | Commentary | Guidelines for a hospital-based AAI program, which has been effective for a hospital in San Paolo, Italy | Did not include number of papers formally reviewed | N/A | N/A | AAI programs can be properly implemented in hospitals if strict attention is paid to animal inclusion criteria and infection control. |
| (Lefebvre et al., 2008a) | Commentary | Provides standard guidelines for animal-assisted interventions in health care facilities, considering the available evidence. | Did not include number of papers formally reviewed | N/A | N/A | Created strict guidelines, centered on evidenced-based literature, for AAI programs to reduce risk of colonization and transmission of hospital-associated infections for the animals and people. |
| (Disalvo et al., 2006) | Commentary | Compared guidelines for therapy animals in hospitals to guidelines for service dogs and family pet visitation | Did not include number of papers formally reviewed | N/A | N/A | Argued that therapy animals should have strict guidelines to reduce adverse events such as phobias, allergies, and zoonotic diseases. |
| (Sehulster & Chinn, 2003) | Review | Centralized CDC guidelines for environmental infection-control strategies and engineering controls to effectively prevent nosocomial infections in healthcare fields. | Did not include number of papers formally reviewed | N/A | N/A | Discussed general infection control policies, but also included therapy animal programs. Recommended minimizing contact with animal bodily fluids, and implementing hand hygiene after each contact. Recommended careful selection of therapy animals and bathing to reduce allergens. |
| (Brodie et al., 2002) | Review | Review of current literature focusing on health risk to patients | Did not include number of papers formally reviewed | N/A | N/A | Zoonoses, allergies and bites - the three issues surrounding pet therapy causing greatest concern - have the potential to be controlled in a supervised health care setting, and can be minimized by taking simple measures. |
| (Guay, 2001) | Review | Review of the most common zoonotic infections that might be expected in the long-term care setting from AAI, with recommendations for prevention and control. | Did not include number of papers formally reviewed | N/A | N/A | Recommends infection control policies and procedures, geared toward management and prevention of the different zoonotic illnesses discussed, should be developed and implemented in all nursing homes offering pet-assisted therapy. |
| (Khan & Farrag, 2000) | Commentary | Critique of current animal therapy programs guidelines in the context of hazard reduction | Did not include number of papers formally reviewed | N/A | N/A | If put into place properly, animal therapy programs can have significant benefit to patients, with minimal risk of animal associated health hazards. |

**Figure 1.**
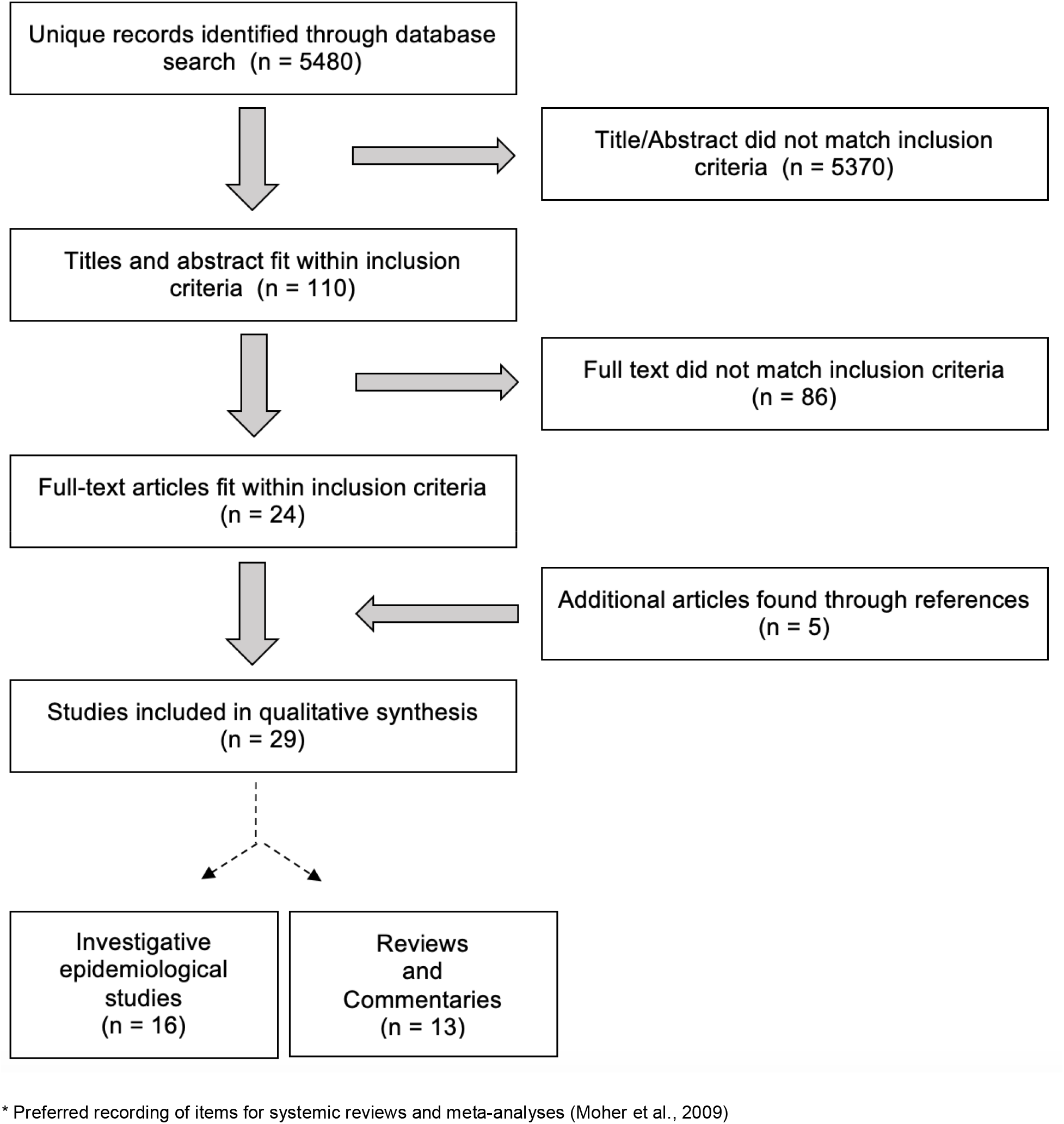
PRISMA* Flow Diagram for Search Strategy. * Preferred recording of items for systemic reviews and meta-analyses (Moher et al., 2009)

### 3.2 Commentaries and Review Articles

Of the 13 commentaries and reviews, there were a total of 7 commentaries and letters to the editors and 6 systematic or unstructured literature review articles. Four of the six reviews (Brodie et al., 2002; Cimolai, 2015; Guay, 2001; Sehulster & Chinn, 2003) and four of the seven commentaries (Disalvo et al., 2006; Khan & Farrag, 2000; Lefebvre et al., 2008a; Murthy et al., 2015) focused on risks associated with infection control. The remaining articles primarily discussed AAI benefits, with only a brief mention of hazard reduction. Zoonotic infection and pathogen transmission were the primary hazards discussed, although some papers mentioned injury risk. One article, endorsed by the Society for Healthcare Epidemiology of America (SHEA), is the current source for the medical community on general guidance for animals in healthcare settings, both summarizing existing policies in hospitals and recommending practical directives to minimize risk (Murthy et al., 2015). In this article, the authors also acknowledge that this field remains insufficiently studied (Murthy et al., 2015). There was a consensus among the reviews and commentaries that with proper hospital infection control protocols in place, the risks associated with animal-assisted activities are minimized. All articles recommended using standardized regulations across healthcare facilities for infection control practices for patients and therapy animals. Three of the articles strongly recommended utilizing expert consultation in various animal and human health care fields, as well as environmental microbiology, to evaluate all possible routes of pathogen transmission (Chalmers & Dell, 2016; Disalvo et al., 2006; Waltner-Toews, 1993).

### 3.3 Epidemiological Studies

The three studies that surveyed hospital infection control policies demonstrated dissimilarities across hospitals. Among the combined 186 facilities surveyed, infection control policies regarding therapy animals varied, with 13% (Linder et al., 2017; Murthy et al., 2015) to 90% (Waltner-Toews, 1993) of healthcare facilities having no existing standardized policies. Only 28% of facilities required documentation that the animal was healthy, and only 29% allowed solely registered therapy animals (Linder et al., 2017). In addition to clinical practice policy discrepancies, animal handler knowledge of infectious disease concerns and adherence to infection control policies varied across and within institutions. Lefebvre et al. found that 20% of 90 surveyed handlers did not practice any infection control and 40% of these handlers were unable to name one zoonotic disease or pathogen that may be transmitted by means of their dog, while Boyle et. al. found that 70% of their 40 handler respondents expressed no concerns regarding infectious disease transmission in AAI settings (Boyle et al., 2019; Lefebvre et al., 2006b). These institutional and individual discrepancies in AAI programs drive diversity in infection control practices both across and within healthcare facilities.

Three studies reviewed electronic medical records to compare a change in the rate of diagnosed infections from AAI exposure. One study evaluated hospital-wide infection rates one year after the introduction of an AAI program in a pediatric hospital and, comparing these rates to the previous year, found no changes in overall infections or detected pathogens reported by the hospital’s infection control committee (Caprilli & Messeri, 2006). Another prospective cohort study followed 11 adult cardiac patients after receiving multiple AAI therapy sessions (average of 13 visits) and found no reports of infection in participants observed during the study period, but did not compare the AAI participants to a control group (Snipelisky et al., 2016). However, another electronic medical record review study identified eight newly-acquired infections two weeks post AAI therapy in nineteen pediatric oncology patients, but could not definitively attribute these infections to the therapy animal visit as there was no control group of hospitalized pediatric oncology patients not receiving AAI therapy (Chubak et al., 2017).

The ten investigative epidemiological studies described cases of either animals or human patients becoming contaminated as a result of an AAI visit. The strongest weight of evidence was from prospective cohort studies in therapy animals (three studies, see **Table 1**). Among these studies, the largest sample size was 200 therapy dogs, with most studies ranging from 10 to 20. In addition, the same group of investigators conducted most of these studies and utilized the same cohort of therapy dogs (Lefebvre et al., 2006a, 2008b, 2009, 2006c; Lefebvre & Weese, 2009). These studies focused on zoonotic pathogen carriage in therapy animals, and detailed cross-sectional prevalence and longitudinal incidence. They observed asymptomatic carriage of both hospital-associated and novel pathogens, such as methicillin-resistant *Staphylococcus aureus (MRSA), Clostridium/Clostridioides difficile, Salmonella, Pasteurella*, and intestinal helminths. This investigator group sampled therapy animals longitudinally over 12 months, and detected incidence rate ratios for therapy dogs with hospital exposure compared to no hospital exposure of 4.7 for MRSA acquisition and 2.4 for *C. difficile* acquisition (Lefebvre et al., 2009). They also identified risk factors for acquiring or being colonized with these pathogens, such as a raw meat diet, being fed treats by patients, and licking patients. One of these studies uniquely sampled therapy animals’ human handlers for hospital-associated pathogen contamination before and after an AAI visit and demonstrated no contamination related to the AAI visit on the handlers’ hands (N=26) (Lefebvre & Weese, 2009). The five other epidemiological studies, not from that investigator group and study population, surveilled therapy animals and found a positive association between therapy visits and zoonotic pathogens. Two were case reports of zoonotic pathogens found in therapy animals (Enoch et al., 2005; Sillery et al., 2004). The three cohort studies found prevalence rates of zoonotic pathogen carriage in therapy animals of 11.8% (Boyle et al., 2019), 18.2% (Coughlan et al., 2010), and 24.3% (Gerardi et al., 2018).

Unfortunately, all of these studies ignored assessment of the human patient, as well as assessment of other individuals involved in AAI, such as healthcare workers, visitors, and, with the exception of the one study mentioned above, the therapy animal handlers. No studies evaluated the hospital environment as a source of pathogens, and the literature included scant data on the clinical health outcomes of the animals themselves. Furthermore, no studies systematically measured risk other than zoonotic pathogens/infectious diseases, such as phobias, allergies, or injuries.

## 4. Discussion

While most of the literature currently available on animal-assisted interventions centers mainly on positive human psychosocial outcomes, there is an apparent lack of information and guiding data surrounding the potential infection control challenges to the inclusion of therapy animals in a healthcare setting. As evidenced by the relatively few and mostly small epidemiological studies discussed in this review (n=10), therapy animals can harbor hospital-associated pathogens, and while not validated in controlled research, these data are consistent with the hypothesis that animal contact with patient populations may increase the animal’s risk for contamination with pathogens. This is best evident in the study that showed therapy dogs that visit hospitals have almost five times higher odds of carrying MRSA as therapy dogs who visit other locations, such as schools (Lefebvre et al., 2009). Additional research is needed to investigate whether therapy animals can serve as pathogen vectors, from being contaminated by contact with one patient, and then transmitting these pathogens to another patient, leading to pathogen exchange. This is critical to test since many patients served by these therapy animals have a compromised health status and may be at higher risk of infection compared to the general population.

While there are proposed guidelines published for AAI in hospitals, senior care facilities, and for individual animal therapy organizations, there are significant differences in infection control policies across these groups (Serpell et al., 2020). This can cause confusion among therapy animal handlers and individuals who participate in AAI programs and may be complicated by a lack of standardized, evidence-based standard-of-care protocols that can be universally adopted. Current guidelines, including the SHEA guidelines, are based on biological plausibility and originate from hospital fomite research and zoonotic transmission in other situations (pets in the home, etc.). Yet it is likely that therapy animals, with their unique exposures and ability to serve as an interactive living fomite, may have microbial communities that are different from standard pet animals. Therefore, exposure to animals in an AAI setting may fundamentally differ from exposure to household pets. This unique exposure profile could logically result in different risk factors and protective factors for pathogen contamination for both participants and the therapy animals. As such, infection control guidelines that rely on previous research on fomites and pet ownership may not realistically reflect adequate control measures for therapy animal exposures.

Our review confirmed an even greater lack of quantitative research on hazards other than infectious disease agents in the context of AAI studies. While some articles commented on the risks of phobias, injuries, negative cultural perception of animals, and allergies, none examined these risk factors empirically. Explanations for few study findings in this area include that these highly-trained animals minimize the potential risk of injury and that patients, along with their supervising medical team, will self-select to participate in these programs, thus reducing therapy animal contact by those patients who have phobias or allergies.

Our review also suggested a lack of effective educational campaigns and open communication networks between hospital infection control departments and therapy animal handlers regarding infection risk. This was suggested both by the variability in control practices among institutions and by the knowledge disparities among handlers observed in multiple studies (Boyle et al., 2019; Lefebvre et al., 2006b; Linder et al., 2017). Without these communication channels, therapy animal handlers may not have a clear understanding of the rationale for infection control protocols, as well as the potential risks towards the patients, their therapy animals, and even the handlers themselves. Continued efforts from infection control departments and hospital program facilitators to provide knowledge-based motivation to adhere to hospital-enacted infection control protocols are essential, directed to both therapy animal handlers and healthcare workers involved in AAI sessions. Without such cohesive collaborations, hospital protocols created for AAI programs can be misinterpreted or poorly executed. In order to minimize the potential risk for all involved, attention should be paid to outreach and education programs that promote safe practices for both therapy animal handlers and hospital staff. In addition to efforts to harmonize infection control regulations across healthcare facilities, individuals involved in AAI should work within the hospital to integrate AAI programs into the overall institutional safety culture in order to maximize the benefits of these programs.

A strong point of the established research is the evaluation of risk factors for pathogen carriage by therapy animals, namely animals fed a raw-food diet and those that have increased interaction with patients (through licking and being fed treats) are more likely to carry zoonotic pathogens. Studies that focus on risk factors can inform interventions to minimize pathogen carriage by therapy animals, and potentially decrease transmission to the patients with whom they subsequently interact. Expanding this work to studies that examine patient-level risk factors (such as concurrent disease conditions or specific animal-contact behaviors) or AAI-level risk factors (such as the number of patients interacting with the animal) will additionally inform the safety practices of these programs and have significant clinical impact. Clear hospital communication channels that impart infection control guidelines, backed by robust evidence-based science on potential risk factors, can empower healthcare workers and handlers to identify and minimize behaviors that pose risk to patients, therapy animals, and themselves.

The most significant knowledge gap is the lack of epidemiological data demonstrating or testing the transmission of zoonotic and hospital-associated pathogens related to AAI therapy sessions. The few published studies have small sample sizes (only two studies included more than 100 animals) and limited longitudinal data (only four retrospective or prospective cohort studies, two from the same cohort). This clearly limits statistical power to demonstrate even associations between pathogen carriage and AAI visits, much less actual illnesses associated with such carriage. Other than those three cohorts, most studies were cross-sectional or case reports, which limits causal inference because of their inherent inability to establish temporality, control for confounding, or account for interpersonal variability. The data from these cross-sectional studies and case reports, therefore, have minimal weight in our understanding of how AAI exposure may relate to pathogen carriage in therapy animals, patients, healthcare workers, and the hospital environment.

At present, the studies that have assessed microbial sharing during a therapy session focused only on the microbial carriage of the therapy animal. Testing only the therapy animal demonstrates carriage of a zoonotic pathogen at a single time point, and does not capture a transmission event. Data and evidence for transmission between patients, animals, and the environment are limited without sampling of all these components. Identification of a transmission event requires longitudinal multi-source sampling (humans, animals, and the environment) with molecular typing to identify and distinguish specific microorganisms. Such data are required to trace the source, pathway, and directionality among therapy animals, the hospital environment, and all individuals involved, including patients, visitors, healthcare workers, and therapy animal handlers.

Longitudinal sampling will also allow insight into whether microbial exposure and transient contamination from AAI conditions can progress into stable bacterial replication and colonization, and then progress to a possible infection in both individuals and therapy animals. In the context of hospital-associated pathogens, it is established that exposure is necessary, but not always sufficient, to progress to infection (Weber & Rutala, 2013); longitudinal sampling can capture these stages of progression, and identify risk factors that promote such progression. This is particularly relevant to clinical outcomes in AAI patient participants, many of whom are children or have compromised health status, making pathogen exposure more likely to progress to an infection. Longitudinal sampling of the therapy animal will also test whether these animals can serve as a vector of disease within and between different hospitals, and in the greater community outside of the hospital, as well as evaluate health outcomes in the animals themselves. With only a few published studies conducted in a small number of single hospitals, and often including the same cohort, the present data are clearly of limited generalizability to other populations.

## 5. Conclusions

Future work in this area should aim to investigate the potential hazards that can occur during a therapy visit, both in terms of potential injury and infection control, and seek to quantify these possible associated hazards, while confirming these hazards do not interfere with the known benefits of AAI. It is recommended that future studies employ a One Health framework, a systems-thinking approach that addresses concerns at the nexus of human health, animal health, and the health of their shared environment, paying particular attention to the relationship between the entities rather than looking at them in isolation (Destoumieux-Garzon et al., 2018). This framework may facilitate future investigations and provide a more holistic view of the microbial dynamics between therapy animals, hospital patients, and the hospital environment. While further research into risk identification is necessary, clinicians and other healthcare workers who implement or are debating implementing an AAI program must also consider their hospital and patient needs, given the clear and established benefits of these adjunctive programs. A rational decision process involves a cost/benefit risk assessment that provides insight into the likely consequences of a proposed action. Balanced with this is the concept of the precautionary principle, which states that without a risk assessment involving hazard identification and analysis, one should minimize exposure to the potential risk. In the case of AAI programs, while there is an ongoing need for corroborating research, the recommended guidelines for animals in the healthcare setting can provide a starting point and scaffold for infection control policies that, when properly applied and followed, have potential to minimize the known and unknown risk factors, while still maintaining the known benefits as an adjunctive patient therapy, with the ultimate goal of making AAI more accessible and sustainable for patients. Promotion of judiciously-executed AAI programs will increase attention to its usage as a complementary therapy, and prompt awareness of the need for further insight into its safety and value as a critical tool for patient benefit.

## Data Availability

The data for this review is accessible on scientific literature databases.

## Author Contributions

Conceptualization – all authors

Methodology – KRD

Data curation – KRD + KBW

Formal analysis – KRD

Supervision – MFD

Writing (original draft) – KRD

Writing (review & editing) – all authors

## Acknowledgements

The authors would like to thank Lori Rosman for her assistance with data collection, as well conceptualization of the project along with Dr. Ellen K. Silbergeld. We would also like to thank Dr. Sharmaine Miller for her consultation.

